# Leveraging implementation science to improve uptake of an ED decision support aid for heart failure patients

**DOI:** 10.1101/2025.11.19.25340621

**Authors:** Katherine A. Vilain, Stacy L. Farr, Danielle M. Olds, Kathryn L. Istas, Maria A. Alvarenga, John A. Spertus

**Affiliations:** Saint Luke’s Hospital, Kansas City, MO; University of Missouri at Kansas City Healthcare Institute for Innovations in Quality; University Health Truman Medical Center, Kansas City, MO

**Keywords:** emergency medicine, heart failure, decision aid, implementation science, qualitative research, quality improvement, evidence-based medicine

## Abstract

**Background:** Changing clinical practice is difficult. An innovative risk stratification process to triage patients presenting to the Emergency Department (ED) with heart failure (HF) exacerbation who were safe for discharge, CodeHF, was inconsistently used in an ED, despite strong evidence of its benefits. Understanding barriers and facilitators to its use is needed to improve adoption. The objective of this study was to understand barriers and facilitators to CodeHF use in routine practice and address determinants of adoption.

**Methods:** A qualitative study was conducted in a four-hospital healthcare system to explore CodeHF implementation and use after three years’ use of CodeHF as part of routine clinical care. The Tailored Implementation in Chronic Diseases (TICD) determinants was used to guide semi-structured interviews and organize themes identified through open coding. Themes were then mapped to Expert Recommendations for Implementing Change (ERIC) strategies.

**Results:** Interviews (n=20) conducted with ED, HF, and hospitalist clinicians; administrative leaders; and researchers revealed five barriers (lack of initial buy-in, validity concerns, lack of clarity around purpose/target, communication gaps, and liability/malpractice concerns) and three facilitators (prompt patient follow-up, decision support, nurse championship). ERIC strategies were adapted to optimize future CodeHF use, and novel strategies were proposed to supplement those from ERIC. Recommendations were created to share with stakeholders and support future implementation efforts.

**Conclusions:** Systematically evaluating the implementation of novel evidence-based treatment strategies can identify barriers and facilitators that can then be mapped to refined implementation strategies. This study exemplifies a pragmatical approach to improve uptake, sustained use, and scalability of an evidence-based intervention for future sites seeking to implement ED pathways for risk-stratifying patients with HF.

**What is known:**

- CodeHF, an algorithmic decision support aid (DSA) for emergency department (ED) physicians triaging disposition for patients experiencing heart failure exacerbation, is underutilized at a large health system despite evidence of clinical and process quality improvements associated with its use.
- Different implementation science tools can be used to assess barriers to uptake of an intervention or to identify potential strategies to improve uptake.

**What the study adds:**

- In addition to barriers found in other ED-based qualitative studies of similar DSAs, this study identifies interdepartmental communication gaps and use of a novel implementation path that ED physicians felt was imposed onto them, hindering initial buy-in to use.
- The study provides a novel example of how to use multiple implementation science tools to plan qualitative interviews, organize coded themes, and tailor strategies to address identified barriers, by mapping identified determinants from the Tailored Implementation in Chronic Diseases framework to known implementation strategies from the Expert Recommendation for Implementing Change (ERIC) framework.
- Five additional strategies are suggested for the ERIC framework: (1) clarify roles and responsibilities, (2) de-implement interventions or intervention components, (3) demonstrate responsiveness to reported barriers and needs, (4) automate processes, and (5) leverage acceptable implementation pathways.

## Introduction

The Hospital Readmissions Reduction Program (HRRP) incentivizes hospitals to reduce heart failure (HF) readmissions.^1^ While many efforts to reduce readmission rates have failed, most have focused on risk stratifying patients at the time of inpatient discharge for closer follow-up.^2–6^ An alternative approach is to more efficiently triage Emergency Department (ED) patients with a history of HF to determine who can be safely discharged home. Using a well-validated risk model that was subsequently used to reduce readmissions, a decision support aid (DSA), CodeHF, was developed and shown to be effective at safely improving discharge rates, with more efficient ED care and fewer 30-day representations.^7,8^ Despite its demonstrated potential to improve the efficiency of HF care in the ED, it was incompletely adopted at a metro area four-hospital healthcare system. During the first 13 months of use, of 1100 eligible encounters, clinicians activated the CodeHF protocol in only 149 (13.5%) cases, with marked variability across sites and physicians.^9^

To better understand why use was low and inconsistent, despite positive process and outcome improvements, a qualitative study of multiple stakeholders was conducted. The study goals were to leverage contemporary implementation science frameworks to identify barriers and facilitators to optimizing CodeHF use. Tailored recommendations were then created to support future CodeHF implementations. This report sought to model methods for developing DSA implementation strategies.

## Methods

### The CodeHF Strategy

The main component of CodeHF is the prospective use of a validated risk score to estimate an ED HF patient’s 7-day mortality.^8,9^ Prior to implementation, ED physicians and HF cardiologists defined a threshold of risk (in this case, a 7-day mortality risk of ≤1%) at which they were comfortable considering discharge from the ED to home, as opposed to hospital admission. Other components of CodeHF for patients discharged home included (1) an educational video for patients and caregivers, and (2) a standardized process for scheduling outpatient cardiology follow-up within 7 days of ED discharge. During its initial implementation, when CodeHF was used, it was associated with a higher rate of ED discharges home, a lower likelihood of those discharged home returning to the ED within 30 days, decreased short stay (<48 hours) admissions, greater proportions of discharged patients having clinic follow-up within 7-days, and shortened times between ED presentation and discharge. Importantly, marked inter-physician variability and low overall CodeHF use was noted.^9^ A better understanding of its inconsistent use was needed to improve care within the health system and to support future use of CodeHF at other institutions.

### Study Design and Setting

To understand why CodeHF use was lower than expected, a qualitative descriptive approach was selected to develop an understanding of the experiences of clinician users and non-users and describe the barriers and facilitators encountered during initial implementation. The study followed Standards for Reporting Qualitative Research (SRQR) reporting guidelines.^10^

Interviews were conducted in person and over video conferencing between November 2021 and January 2023 by a team of investigators trained in qualitative methods, health services research, implementation science, and clinical medicine. This project was designated as Quality Improvement by the local Institutional Review Board and all participants provided verbal consent. This study was funded by the Healthcare Institute for Innovations in Quality, University of Missouri—Kansas City.

The first two participants identified were ED CodeHF physician champions. Targeted and snowball sampling identified additional key personnel for interviews. These included other ED physicians and nurses, cardiologists, hospital leadership, IT staff and leadership, and researchers involved in the initial CodeHF implementation. In addition to transcribed interviews, interview notes, and email messages between the researchers and three interviewed ED physicians were included as source material.

### Theoretical Framework

The Tailored Implementation for Chronic Diseases (TICD) determinants framework was used to guide the creation of semi-structured interview guides and organize the qualitative analyses.^11^ The TICD is a framework for identifying and classifying determinants - barriers and facilitators - to implementing interventions for chronic disease care. It has seven domains: individual health professional factors; patient factors; professional interactions; incentives and resources; capacity for organizational change; social, political, and legal factors; and guideline characteristics.

### Interview Procedures

Interview questions were based on selected TICD domains to ensure all potential barriers and facilitators were probed among individuals who participated in implementing or were intended end users of the tool. Interview guides were customized to four groups: clinician users (e.g., ED physicians and nurses) and nonusers (e.g., hospitalists and HF specialists who received admitted patients after ED triage); IT members who programmed the order set and created usage reports; hospital administrators; and the researchers involved in the original implementation. Interview guides were developed over several iterations among the qualitative research team. After the first interview, minor modifications were made to wording and question order to optimize interview flow. A common set of interview items is shown in Table 1 paired with relevant TICD domains. Interviews were recorded digitally, with additional digital notes taken. No financial incentives were provided to participants. Interviews were conducted until multiple participants from each of the four groups were included and thematic saturation was achieved.

**Table 1.** Examples of TICD domains & determinants paired to interview guide items, by interviewee role in CodeHF.

| TICD Domain | Determinant* | Example Interview questions |  |
| --- | --- | --- | --- |
|  |  | Clinician/Administrator | IT implementor |
| Recommendation | Source/quality/strength of evidence supporting the recommendation | What do you think about the evidence supporting the CodeHF program? |  |
|  | Cultural appropriateness | What is your perception of the safety or appropriateness of CodeHF? | How does CodeHF align (or not align) with your prior experience implementing interventions that require an EHR change? |
|  | Consistency with other guidelines | Are the components embedded in the CodeHF project consistent with the goals and quality metrics you work with routinely? |  |
|  | Compatibility w/workflow | How compatible is CodeHF with your current clinical workflow? | How similar are the components embedded in the CodeHF pathway to other order sets and quality metrics you work with routinely? |
|  | Effort | How much time and effort is required to use the CodeHF Program? | How much time and effort has been required on your part to work on CodeHF (e.g., EHR build, maintaining/ updating/ modifying)? How has that changed over time? |
| Individual Professional | Awareness and familiarity with the recommendation | What do you know about the CodeHF program? Describe the education you have received about CodeHF. How could this be improved? |  |
|  | Agreement with the recommendation | Are the goals of CodeHF realistic? Feasible? |  |
|  | Expected outcome | What is your understanding of the goals or anticipated outcomes of CodeHF? |  |
|  | Intention and motivation | What incentives motivate you to adopt a practice or change your behavior? |  |
|  | Self-efficacy | How will (/how does) using CodeHF impact your ability to do your job, and your efficiency? | How did decision-making about the EHR changes for CodeHF impact your efficiency? |
|  | Capacity to plan change | Do you/your team have the capacity to make changes that would improve use of CodeHF? |  |
|  | Self-monitoring and feedback | How are you able to review your own CodeHF use and patient outcomes? How does that affect your use of CodeHF? | What do you know about how much CodeHF is being activated in your ED? |
| Patient | Patient needs | How does CodeHF address or not address patients' needs? |  |
|  | Patient preferences | How do provider preferences correspond to actual patient values and preferences? |  |
|  | Patient motivation | How [well] do patients and/or their caregivers participate in CodeHF education? |  |
| Professional Interactions | Communication and influence | How often and in what ways does your team communicate about CodeHF? Describe your colleagues' views of the CodeHF Program. How does this align with your views? How has this influenced your use of CodeHF? | How often and in what ways does your team communicate about CodeHF? Describe your colleagues' views of the CodeHF Program. How does this align with your views? What do you know about how Code HF is being received by end users? |
|  | Team processes (skills) | Are there team or workflow issues that affect your use of CodeHF? | Are there team or workflow issues that affect end use of CodeHF? |
| Incentives and resources | Availability of necessary resources | What resources would be helpful in standardizing the use of CodeHF in your department? |  |
|  | Financial incentives & disincentives | What are the financial incentives available to you and your colleagues for use of recommended practices/strategies? |  |
|  | Nonfinancial incentives & disincentives | What are the non-financial incentives? |  |
|  | Information system | How does the electronic health record system facilitate or hinder your use of CodeHF? How could this be improved? | Are there technical issues that affect end use of CodeHF? |
|  | Assistance for clinicians, including decision aid/support, clinical supervision | Do you or your colleagues have the resources needed to implement CodeHF? (checklists, patient information, decision aids, decision support, clinical supervision) |  |
| Capacity for organizational change | Mandate, authority, accountability | How do internal and external rules, regulations, policies or procedures help or hinder use of CodeHF? | Who participates in decisions about implementing CodeHF? |
|  | Capable leadership | What changes require leadership or management? Do they leaders/managers have the capacity to execute the change(s)? |  |
|  | Regulations, rules, policies | How do organizational guidelines/policies support (hinder) the adoption of CodeHF? Specify/example. |  |
|  | Priority of necessary change | How much of a priority is the use of CodeHF, compared with other initiatives and activities going on in your setting? |  |
|  | Monitoring & feedback | How useful is monitoring and feedback? Is it available? | How do you receive feedback about changes you have made to the EHR? |
| Social, political and legal factors | Payer/funder policies | Are there any payer or funder policies or issues that you're aware of that either help or hinder CodeHF? |  |
|  | Malpractice liability (real & perceived) | Do real or perceived risks of malpractice complaints facilitate or hinder implementation of necessary changes? |  |

### Data Analysis

Interview recordings were transcribed by Rev.com and de-identified for analysis within Dedoose software.^12^ Open coding was used to explore CodeHF barriers and facilitators, and a codebook based on emergent findings was developed by four members of the research team who reviewed interview transcripts. Two members of the coding team independently coded each interview, with discrepancies resolved through group discussions with all coders. Codes were grouped into categories of factors that facilitated or inhibited CodeHF use. These were shared with and approved by a subset of interviewees as a form of member checking. Categories were mapped to TICD domains.

### Implementation Strategy Mapping and Tailoring Recommendations

The overall study process is illustrated in Figure 1. The Expert Recommendations for Implementing Change (ERIC) summarizes implementation strategies from published literature.^13^ In the final phase of this study, ERIC strategies were examined for potential alignment within each TICD category and the identified determinants. Additional strategies were also identified, as needed. After determinants were mapped to strategies, tailored recommendations were generated in the context of planning for CodeHF re-implementation at the study site and to guide future implementation at other institutions.

**Figure 1.**
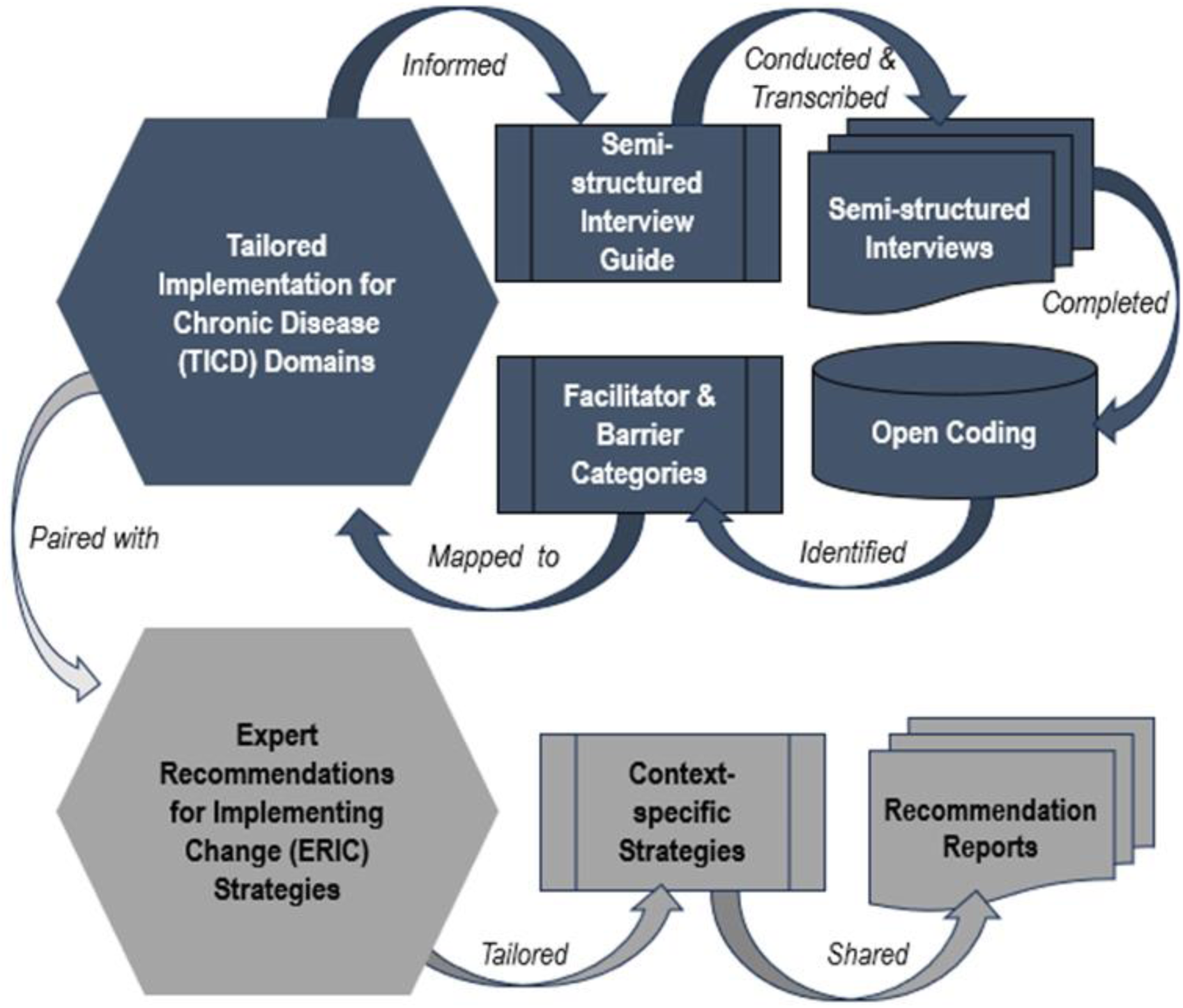
Study Process Map. The figure illustrates the overall study process with phase chronologically linked using arrows, and colors reflecting the implementation framework(s) informing the phase. The process begins with the Tailored Implementation for Chronic Diseases (TICD) Domains and moves to the development of the semi-structured interview guides, proceeding through the dark blue icons, where the processes and products were informed by the TICD. After mapping facilitator and barrier categories identified through open coding back to the TICD, they were paired with (mapped to) Expert Recommendations for Implementing Change (ERIC) Strategies. The lighter grey icons are informed by convergent use of the TICD and ERIC. Hexagons represent implementation science frameworks. The predesigned process icon (rectangle with vertical borders) represents products or outcomes of a process. The multidocument icon represents processes or products of a similar type. The magnetic disc icon represents a process.

## Results

### Participants

Twenty interviews were conducted with 24 participants – 17 individual semi-structured interviews, and 3 focus groups with 2, 2, and 3 participants each (Table 2). Participants were 64% male. Ten were physicians (5 ED, 1 internal medicine, and 4 cardiologists); four were nurses (1 ED, 1 cardiology, and 2 nurse educators). The remaining eight represented IT (3) and hospital (2) leadership, along with researchers (3).

**Table 2.** Interviewee characteristics.

| **Characteristic** | **Respondents, No (%)** |
| --- | --- |
| Total participants | 22 (100%) |
| Male | 14 (64%) |
| Female | 8 (36%) |
| Stakeholder Type |  |
| Physicians | 10 (45.4%) |
| Emergency Medicine | 5 (22.7%) |
| Internal Medicine | 1 (4.5%) |
| Cardiologists | 4 (18.2%) |
| Nursing | 4 (18.2%) |
| Emergency Medicine | 1 (4.5%) |
| Cardiology | 1 (4.5%) |
| Education | 2 (9.1%) |
| IT Leadership | 3 (13.6%) |
| Researchers | 3 (13.6%) |
| Hospital Leadership | 2 (9.1%) |

### Themes

Coding revealed eight major factors associated with barriers and facilitators to optimal CodeHF use. Mapping to TICD domains revealed at least one example from each major domain.

### Facilitators

Three facilitators were identified, including the seven-day follow-up appointment, ED throughput, and the role of nurses in the workflow. Table 3 shows example quotes for each theme, along with the TICD domain and determinant deemed most closely associated with each theme.

**Table 3.** Facilitator Themes and Supporting Quotes with Alignment to TICD Determinants.

| # | QUOTE | TICD Domain and Determinant |
| --- | --- | --- |
| **Facilitator Theme 1. Seven-Day Follow-up Appointment** |  |  |
| F2.1 | *"...[I]t did speak to one of the things that makes [ER physicians] very anxious, which is, 'I don't want to send someone home unless I know they've got follow-up,' and these patients got follow-up."* | **Professional Interactions:** Referral Processes |
| F2.2 | *"[I]f we send them home, we have the benefit of making sure that we can get...arranged follow-up for these patients either through a video or in-person visit with cardiology."* | **Professional Interactions:** Referral Processes |
| F2.3 | *"We did hear some buy-in. A lot of the ED physicians we talked with, like the idea of knowing that the patients would follow up with a cardiologist, after they sent them home within 7 days. That's huge."* | **Professional Interactions:** Referral Processes; **Patient factors:** Needs |
| **Facilitator Theme 2: ED throughput** |  |  |
| F1.1 | *"The whole idea that if we get people started off on the right foot in the ED and get them into the hospital, at a better starting point, they get out faster. If throughput is better, beds become more available. Everybody wins. I think framing this as part of a system of care that gets people through the system faster is always good."* | **Incentives & Resources:** Availability of necessary resources |
| F1.2 | *"I want to make sure that we benefit the patients and that if we feel like we can send them home, we can send them home and not tie up a bed that's maybe not necessary... We want to do right for the patient, but we also need to have beds for our patients."* | **Incentives & Resources:** Availability of necessary resources |
| **Facilitator Theme 3: Role and Attitude of Nurses** |  |  |
| F3.1 | *"I don't think it affects [our workflow] at all... We kind of work off of tasks and things that we have to do and that just is another task that we do."* | **Individual Healthcare Professional:** Cognition (attitudes towards guidelines in general, intentions and motivations); **Recommendation:** Recommended behavior (compatibility, consistency with other guidelines) |
| F3.2 | *"With something significant, like a code heart failure, I'm a fan of BPAs, best practice alerts, because it just puts it in your face. It's a good reminder."* | **Individual Healthcare Provider:** Cognition (attitudes towards guidelines in general, intentions and motivations); Behavior (self-monitoring) |
| F3.3 | *"We have a great team. We really do, with doctors, with nursing staff, with techs. I mean, we rely on each other so much. It's really... It's great. Can't say that with every place, but it's great down in the* | **Professional Interactions:** Team processes |
|  | *emergency department."* |  |

#### Facilitator Theme 1: Seven-Day Follow-up Appointments

The most-cited facilitator across all interviews was the establishment of a new process whereby patients discharged using the CodeHF DSA are contacted within 48 hours to schedule an outpatient cardiology follow-up visit within 7 days. Interviewees universally appreciated this aspect of the program and felt that it directly addressed a concern of discharging patients to home from the ED.

#### Facilitator Theme 2: ED Throughput

While initial concerns that the CodeHF program would be time consuming, early analyses of the pilot project demonstrated faster ED ‘door-in to door-out’ time. Faster ED throughput was perceived as a very positive impact from ED physicians’ perspective, even among CodeHF ‘non-users’. This benefit was potentially undermined by the trade-off that while overall ED time was reduced, CodeHF required more physician-patient communication. Both concepts were mentioned by one ED physician user: “I think it does reduce the actual time. It increases the time, I’m guessing, that doctors have to spend in the room.”

#### Facilitator Theme 3: Role of Nurses in CodeHF

In contrast to the perceptions of many ED physicians, ED nurses generally showed enthusiasm around the potential benefits of CodeHF on processes and patients. They found that CodeHF aligned with similar initiatives that they valued and cited no difficulties or misgivings about incorporating CodeHF into their workflow. ED nurses strongly valued anything that helped their patients, and accepted physician leadership on what was most appropriate.

### Barriers

Identified barriers included ownership of and buy-in to the CodeHF project, concerns about scientific validity of the mortality risk score and evidence of effectiveness of the pathway, technological errors and limitations, communication gaps, and liability/malpractice concerns. Table 4 shows example quotes for each theme, along with the TICD domain and determinant deemed most closely associated with that theme.

**Table 4.** Barrier Themes and Supporting Quotes Aligned to TICD Determinants.

| # | QUOTE | TICD Domain and Determinant |
| --- | --- | --- |
| **Barrier Theme 1: Project Buy-in and Ownership** |  |  |
| B1.1 | *"You need more champions, and you need more people that are willing to stand up in front of their own peers and not a specialist that's coming to them. To say, this is why I believe in this."* | **Capacity for Organizational Change:**<br>Authority, accountability |
| B1.2 | *"The ED has a special way of doing things too, yeah. Even one physician isn't enough, they have to debate things internally on their own. And every group has some different ways that they come together and that they would prefer to vet. You know what I mean? And the ED likes to do that, and they like to debate each other. And I don't think that there was enough support for this one."* | **Individual Health Professional Factors:**<br>Professional Behavior;<br>**Capacity for Organizational Change:**<br>Relative strength of supporters and opponents |
| B1.3 | *"...[P]art of the perception is that it is very odd that the push to abide by this comes from research and not from practicing clinicians. It feels like it is coming from a non-clinical department."* | **Capacity for Organizational Change:**<br>Authority, accountability |
| B1.4 | *"You needed more vetting and more people to believe in the mission prior to trying to push the mission on someone."* | **Incentives & Resources:**<br>Quality assurance systems & assistance for adherence |
| B1.5 | *"If you want this adopted, you need to get every single [inpatient] cardiologist on board. And hospitalist."* | **Professional Interactions:**<br>Communication & influence |
| **Barrier Theme 2: Validity of the score** |  |  |
| B2.1 | *"I think there's a lot of confusion and limitations to the score. I think that there's uncertainty to the physicians on how valid that score is, and specifically in an ED setting, are these valid and reproducible and how accepted is this patient's Heart Failure Score?"* | **Guideline Factors:**<br>Quality & clarity of evidence supporting the recommendation;<br>**Individual Healthcare Professional:** Cognition – expected outcome |
| B2.2 | *"EPIC has a major limitation on how we document what their oxygen delivery is, whether they're on room air or cannula or face mask, or what. And so the patient may always have an oxygen saturation in the nineties or mid-nineties, and that's all the score's extracting."* | **Incentives & Resources:**<br>Information System |
| B2.3 | *"There's so much gestalt, I think, that comes in dealing with a heart failure patient. And so I may be seeing a patient that I think needs to come into the hospital because they're having an oxygen requirement, or for one reason or another, they got new onset heart failure, or something like that, and I might do the score and the risk comes back at 0.87, and it's telling me this person is safe for discharge. And I said, 'No. As a physician, I'm seeing this patient. I disagree. I'm going to admit that patient.' And I might have a patient* | **Individual Health Professional Factors:**<br>Knowledge and Skills (Domain knowledge);<br>Cognitions (Agreement with the recommendation, self-efficacy) |
|  | *who looks great, I diuresed him, they've got chronic heart failure, and I do the run, the score, and it comes back at 3.47 saying that my person's got an increased 30-day mortality. And I say, 'Eh, no, this person looks great. I'm going to let him go home and follow up as an outpatient.'"* |  |
| B2.4 | *"I think that there's uncertainty on how valid that score is, and specifically in an ED setting, are these valid and reproducible?"; "...[I]t was developed and validated at a single site and without second-site validation, in Canada."* | **Guideline Factors:**<br>Strength, Source of recommendation |
| **Barrier Theme 3: Technology Errors and Limitations** |  |  |
| B3.1 | *"The compliance data that we have received for enacting code heart failure for almost two years now has been 0%, 0%, 0%, 0%. And then you've got the providers saying, "We're doing this. How could it be 0%?"; "And I think that they tried to do it and [they realized] 'we're not getting credit for it. This data doesn't align with what we know we're doing.'"* | **Incentives & Resources:**<br>Quality assurance systems; **Individual Healthcare Professional:** Self-monitoring or feedback |
| B3.2 | *"[The time it takes for the labs to come back could be a barrier to running the score], just because if something's a task, you want to get it over with and so if you can't do it now, then some people will just be like, 'Well, I'll get to it later,' but then they never get to it. So not having those labs back immediately would maybe give them some time to forget about it."* | **Incentives & Resources:**<br>Information system; **Individual Healthcare Professional:** Nature of the behavior |
| **Barrier Theme 4: Communication Gaps** |  |  |
| B4.1 | *"... [I]t would be frustrating if you're relying on implementing something that came out of the evidence-based practice of cardiology and you're talking to a cardiologist who has hopefully been given the same opportunity to learn about this as you. And they are the ones who say 'I have no idea what you're talking about.' Of course the ED doc is going to be like, 'I don't even know why we do this.'"* | Professional Interactions:<br>Communication & influence; Individual Healthcare Professional: Awareness and familiarity with the recommendation |
| B4.2 | *"...[T]he communication between [the different system hospitals], everything was kind of so piecemeal, because I had champions at each place, but I needed somebody that's controlling from above, from top down, so that intervention goes to its all sides."* | **Incentives & Resources:**<br>Regulations, Policy; Capacity for Organizational Change: Authority, Accountability |
| B4.3 | *"And the other thing to remember, within the last six months, I tried and tried and tried to get ED and cardiology to come together and have a discussion about this. And I could never get anybody to commit to a time to come in and talk about it."* | Professional Interactions:<br>Communication & influence |
| **Barrier Theme 5: Concerns about Liability & Malpractice** |  |  |
| B5.1 | *"I'm sitting there defending a case to that trial attorney, that I sent home a patient that then decompensated and died, I say, 'But my* | **Social, Political & Legal:**<br>Malpractice |
|  | *[CodeHF] score was 0.8, and it said that I was okay to send him home.' And they say, 'But doctor, I don't know what the hell that thing is.' You know, 'There's a couple of white papers on it, but did you not use your clinical acumen and decide that this patient needed to come into the hospital?'"* |  |
| B5.2 | *"... [S]ome of our lawyers have asked us, 'Is this part of the legal medical record? Yes or no?' And yes, it was... this provider was presented with this lab value ... and they should have taken this action, and they didn't. And prosecuting attorneys are trying to now sue that provider."* | **Social, Political & Legal:**<br>Malpractice |
| B5.3 | *"...[F]rom a regulatory standpoint, if you got a standing order and you don't use it and Joint Commission finds out about that, things aren't going to go well for you."* | **Capacity for Organizational Change:**<br>Regulations, policies |

#### Barrier Theme 1: Ownership & Buy-in

The foundational barrier to optimal CodeHF use was a low sense of ownership by ED physicians, based on distrust around its origins as a QI project. ED physician investigators ushered CodeHF implementation through a novel pathway that allowed any hospital staff member to apply for QI project support, which circumvented the usual processes of project advocacy by the ED’s Evidence-based Practice Team (EPT). While the EPT agreed to implement the project, there was resistance by its members because it did not originate from within the EPT committee.

#### Barrier Theme 2: Validity of the Score

Some ED physicians expressed skepticism about the CodeHF evidence base. Concerns around validity included the comprehensiveness or accuracy of the risk score, which at the time did not incorporate results of a blood test commonly used in the U.S. for heart failure (i.e., B-type natriuretic peptide [BNP]); inappropriateness of use in patients with alternative potential diagnoses; the small size and design of the original health system pilot study; and the generalizability of a tool developed and validated in Canada. This skepticism was exacerbated when the estimated mortality risk did not align with their clinical assessment. This concern persisted even after efforts to highlight that the score was intended to supplement, not replace, clinical judgement.

#### Barrier Theme 3: Technology Errors and Limitations

A part of the implementation strategy was to provide periodic reports of CodeHF use and outcomes. However, errors in the feedback reports occurred and users felt they were not getting appropriate credit for using CodeHF. In addition, limitations of the electronic heath record precluded lab values automatically populating the mortality risk calculator, requiring additional steps for manual data input.

#### Barrier Theme 4: Communication Gaps

Cross-departmental communication gaps compounded the initial reticence for using CodeHF. During the initial implementation of CodeHF the focus was on ED physicians, rather than on those receiving admitted patients. Unfamiliarity with the risk score by admitting hospitalists and cardiologists gave ED physicians the perception that it lacked value, particularly among ED physicians initially skeptical of CodeHF. While subsequent educational efforts outside of the ED were pursued, a lack of centralized communication around the project, and the large number of hospitalists to educate, likely contributed to their lack of familiarity with CodeHF. Another major communication gap, likely underscoring Barrier Theme 2, occurred between the original project initiators and the end users of CodeHF. A researcher stated: “We explained to the group that the CodeHF score should be treated as any other biomarker and merely an additional piece of information, but not the sole deciding factor.”

#### Barrier Theme 5: Concerns about liability & malpractice

ED physicians dissatisfied with CodeHF also cited concerns about potential malpractice risks associated with its use, particularly in the event of a patient death with a score suggesting the patient could be discharged home. In contrast, others felt that documenting a patient’s risk for safe discharge could be helpful in litigation, should an adverse outcome occur.

### Recommendation Strategies

Table 5 provides tailored recommendations targeting each of the identified barriers and facilitators. These were developed with ERIC strategies and supplemental strategies, as appropriate. The third column of the table shows the relevant ERIC strategy to which barriers were mapped, with the proposed additions in **bold** font, while the fourth shows tailored recommendations. Novel strategies proposed include **Leveraging accepted implementation pathways**, **Automating processes**, **Demonstrating responsiveness to reported barriers**, **Deimplementation of unvalued components**, and **Clarifying roles and responsibilities**.

**Table 5.** TICD domains, ERIC strategies, and tailored recommendations by theme.

| Interview Themes | TICD Domains & Determinants | ERIC Strategy (suggested additions in bold) | Tailored Recommendations |
| --- | --- | --- | --- |
| **Barrier Theme 1: Project Ownership & Buy-in** | **Capacity for Organizational Change:** Authority, accountability, Relative strength of supporters and opponents | Use advisory boards and workgroups; Involve executive boards | Obtain broad system support for CodeHF through engagement with hospital C-suite and end users |
|  | **Individual Health Professional Factors:** Professional Behavior | Identify and prepare champions | Identify new clinical champions to lead a reboot of CodeHF |
|  | **Incentives & Resources:** Quality assurance systems & assistance for adherence | Develop a formal implementation blueprint; Provide local technical assistance; Develop and implement tools for quality monitoring | Formally plan for transparency, centralized and regular communication, and processes for rapidly addressing emergent issues |
|  | **Professional Interactions:** Communication & influence | Conduct local consensus discussions; Organize clinician implementation team meetings; **Leverage accepted implementation pathways** | New champions should focus on buy-in from users and implement a reboot through the ED EPT. |
| **Barrier Theme 2: Validity of the score** | **Guideline Factors:** Quality & clarity of recommendation | Purposefully reexamine the implementation | Clarify appropriate use cases for CodeHF and its intended target population |
|  | **Incentives & Resources:** Information System | Change record systems; **Automate processes** | Work with EPIC to automate population of lab values in score calculator |
|  | **Guideline Factors:** Strength of recommendation | Stage implementation scale-up; Conduct educational meetings | Provide local models of successful implementation and outcomes of CodeHF at other sites in the area to encourage (re)adoption. Educate clinicians on updated validation and outcomes data and improvements to the intervention |
| **Barrier Theme 3: Technology Errors and Limitations** | **Incentives & Resources:** Quality assurance systems;<br>**Individual Healthcare Professional:** Self-monitoring or feedback | Develop and implement tools for quality monitoring; Facilitate relay of clinical data to providers; **Demonstrate responsiveness to reported barriers** | Reprogram user compliance reports for accuracy. Build user-friendly interface for accessing easily readable ED- and provider-level reports. Build trust by responding to technical and communication issues promptly |
|  | **Incentives & Resources:** Information system;<br>**Individual Healthcare Professional:** Nature of the behavior | **Deimplement components** | Deimplement educational video component due to technical failure. |
| **Barrier Theme 5: Communication Gaps** | **Professional Interactions:** Communication & influence; **Individual Healthcare Professional:** Awareness and familiarity with the recommendation | Build a coalition | Intentionally include representatives from cardiology, hospitalists, and ED in implementation team and all ongoing meetings and education |
|  | **Incentives & Resources:** Regulations, Policy;<br>**Capacity for Organizational Change:** Authority, Accountability | Visit other sites; Model and simulate change | Encourage systemwide adoption when ED championship at origin hospital is strong |
| **Barrier Theme 5: Concerns about liability & malpractice** | **Social, Political & Legal:** Malpractice | Develop educational materials; **clarify roles & responsibilities** (including legal liability) | Re-educate ED physicians on liability laws; include language in the tool to assure users |
|  | **Capacity for Organizational Change:** Regulations, policies | Mandate change | Reinstate standing order once clinicians accept CodeHF |
| **Facilitator Theme 1: Seven-Day Follow-up Appointment** | **Professional Interactions:** Referral processes; **Patient factors:** Needs | Facilitate relay of clinical data to providers | Promote uptake of CodeHF by emphasizing the benefit of its built-in patient follow-up component |
| **Facilitator Theme 2: ED Throughput** | **Incentives & Resources:** Availability of necessary resources | Increase demand | Promote uptake of CodeHF by emphasizing the benefit of reduced ED time |
| **Facilitator Theme 3: Role and Attitude of Nurses** | **Professional Interactions:** Team Processes | Revise professional roles; Recruit, designate, and train for leadership | Leverage nurse championship as a readily available resource for physicians implementing new practices in the ED. |
|  | **Individual Healthcare Professional:** Cognition (attitudes towards guidelines in general, intentions and motivations);<br>**Recommendation:**<br>Recommended behavior (compatibility, consistency with other guidelines)* | Model and simulate change | Encourage nurse champions to promote use of CodeHF |

Barriers identified in this analysis came from all domains of the TICD, suggesting CodeHF optimization will require simultaneous use of interacting strategies to address the interconnected obstacles.

Adopting multiple strategies may comprehensively address the identified barriers. Facilitators, such as enthusiastic CodeHF support by ED nurses, rapid cardiology follow-up of discharged patients, and its positive impact on ED throughput, should be leveraged. The most critical recommendation for implementing CodeHF is encouraging ED clinicians to lead its design and launch through the approved pathway of bringing evidence to practice (e.g., an ED EPT in this health system).

## Discussion

Changing clinical practice is difficult, particularly in a fast-paced, high-pressure setting, such as the ED. A qualitative evaluation was conducted to understand why an effective protocol had variable adoption by ED physicians to improve re-implementation and support implementation at future sites. By interviewing multiple CodeHF stakeholders, three facilitators to CodeHF use were identified: better outpatient follow-up for discharged HF patients, improved ED throughput, and nursing support. Offsetting these facilitators, five significant barriers were identified: insufficient initial buy-in from the ED; skepticism of clinical evidence supporting the pathway; IT challenges; communication; and liability concerns. Lessons learned can guide re-implementation at this health system and support future sites seeking to improve the efficiency, safety, and patient-centeredness of managing HF exacerbations.

The identified themes generally align with other implementation evaluations of ED provider-facing DSAs. Fujimori et al. (2022) explored the acceptance, barriers, and facilitators of implementing AI-based decision support in the ED,^14^ which highlighted that while these tools are generally well-received, challenges included concerns about over-reliance on algorithms for diagnosis, which was paralleled in the CodeHF study by confusion about the tool’s intended use as a support rather than a replacement for clinical acumen. Similar to CodeHF, a study by Billah and colleagues assessing ED physicians’ determinants to hypothetical use of two patient risk-stratification tools (SynDA for syncope and Chest Pain Choice) included concerns of medicolegal risk, lack of need, and skepticism about the tools’ validity. Facilitators included positive attitudes towards shared decision making and improved patient follow-up.

An important insight was the means by which CodeHF was introduced. Although the project was selected, designed, and led by an ED physician, it did not follow the institution’s established EPT pathway. This barrier may be unique to this implementation as we were unable to find another qualitative analysis of DSA implementation that identified the mode of translating evidence into practice as a barrier. However, there is evidence that shifting implementation leadership to the ED and using their preferred process reflects distributing information and decision-making authority to those most affected by the implementation.^15^ Communication gaps between departments also did not appear to be a theme identified in other published DSA implementation studies.

This study should be interpreted in the context of several potential limitations. Members of the interviewing team work closely with the research staff interviewed. Where possible, the team member who did not have a previous working relationship with the research staff led the focus groups and interviews, but the fact that they were researchers may have biased respondents’ comments. Second, while thematic saturation was achieved, other determinants and themes may have arisen had further probing been conducted or a broader group of participants been included. Third, findings of this study may not be generalizable to other settings. Clinical cultures differ and unique sets of barriers and facilitators might be relevant to different EDs. However, many findings align with similar investigations of barriers to ED support tools.

While this evaluation addresses a unique protocol implementation, the methods are easy to reproduce in other settings. Moreover, by pre-defining implementation framework and strategy lists, this study can model an approach for identifying barriers and facilitators to refine strategies for addressing identified determinants. Specifically, the TICD framework was used as a guide for developing interview questions and for coding categorization, then mapped to ERIC strategies. Similar linking between implementation tools or frameworks has been done in other studies. For example, Waltz describes the development of the Consolidated Framework for Implementation Research (CFIR)-ERIC Implementation Strategy Matching Tool, which involved self-identified implementation experts mapping ERIC strategies to specific CFIR-based barriers.^16^ ERIC is a mutable list, so when no strategy appeared relevant to the CodeHF determinant, new strategies were created, as others have done.^17^ These identifying, mapping, and tailoring-and-creating processes allowed development of a tailored recommendation report for re-implementation with high confidence in its depth and breadth.

## Conclusion

This study demonstrates the utility of implementation science frameworks and tools in tailoring strategies for improving clinical DSA uptake. Use of the TICD to develop interview guides and categorize resultant codes facilitated comprehensive examination of the many sources of facilitators and barriers – structural, cognitive, interpersonal, intervention-specific, etc. -- in interviews. Similarly, the ERIC list was useful for identifying potential strategies for overcoming barriers to revive CodeHF at the study site and to prevent future implementations from repeating the challenges encountered in this experience. Lessons learned from this method of linking the TICD domains to ERIC implementation strategies to create context-specific re-implementation recommendations may be applicable to other health systems seeking to improve the value of healthcare.

## Data Availability

Because transcribed interviews contain potentially identifying information, illustrative quotes in the manuscript have been selected or altered to obscure the speaker's identity. Full interview transcripts are therefore not available.

